# The dangerous of listen it but not understand it: The effect of interoceptive awareness as the underlying mechanism between anxiety and NSSI behaviors

**DOI:** 10.1101/2023.02.26.23286477

**Authors:** Diyang Qu, Yi Feng, Runsen Chen

## Abstract

The underlying mechanism between anxiety and NSSI behaviors remains unclear. Interoceptive awareness has been suggested as a strong transdiagnostic risk factors for emotional disorders. Following the body-mind approach, we thus aimed to examine the potential indirect effect of three important dimensions of interoceptive awareness. The effect of study variables on the lifetime NSSI behaviors was first tested on 5281 Chinese participants, then the indirect effects were further clarified among individual with NSSI behaviors. We found that Emotional awareness and Body listening plays an indirect effect between anxiety symptoms and NSSI frequent, but in a opposite way. Future preventive intervention program in strength individual’s abilities of connection body and emotion has been strongly highlighted.

## Introduction

Non-suicidal self-injury behavior (NSSI) has becoming one of the most serious public health problems over the worldwide nowadays, given its long-lasting effect on individual’s adjustment outcomes, including, higher level of psychosocial dysfunctions (Garisch & Wilson, 2015), psychological symptoms (Lundh et al., 2011), and even a precursor of suicide attempts (Daukantaitė et al., 2021). In this case, the tailored prevention and intervention are urgent to reduce these issues; but it has been limited by the underexplored mechanism underlying the questions, what brings individual practice these maladaptive behaviors.

According to the cognitive emotional model and existing studies, NSSI is commonly performed as an emotion regulation strategy, which help individual decrease their negative emotional experience in short-term (Hasking et al., 2017; Taylor et al., 2018; Wolff et al., 2019). In this case, the robust association has been found between anxiety and NSSI (Bentley et al., 2015). In addition, previous studies further showed that, anxiety is uniquely associated with NSSI behavior than other emotional dysfunction problems (Bock et al., 2021). However, research identifying factors that may explain this pathway is lacking.

In the recent years, the importance of interoceptive awareness has been highlighted. It refers an individual’s ability to identify, access and understand their internal body signals; and further been usefully conceptualized within a dimensional framework (Garfinkel et al., 2015), for example, individual’s noticing and listening to their body and abilities of awareness the connection between body and emotion.

In line with the integrate affective and cognitive processing model of anxiety disorders, individual with higher level of anxiety, may overly sensitive or even ampliative their body signal, in which, leading to more maladaptive adjustment outcomes (Paulus & Stein, 2006). However, to the best of our knowledge, no studies has explored the effect of these potential pathways on individual’s NSSI behaviors. Moreover, to what extent the different dimensions of interoceptive awareness may contributes to this association still remains a large question; in this case, clarifying the mixed pathways would significantly guiding to a more cost-effective preventive intervention program in this “on the edge problem”.

## Methods

### Participants and procedure

The convenience sampling was used, which covered college students in Beijing, China in the year of 2022. A total of 5281 University students (33% were male, N = 1719; and 67% were female, N =3562), M_age_ = 20.8, SD_age_ = 2.66 met inclusion criteria (e.g., passing three out of the four attention check questions) were included in later analysis. Informed consent was obtained from all participants on the information page, before they starting to filling out the questionnaire. The ethical approval was obtained from the Ethics Committee of Central University of Finance and Economics, Beijing, China.

### Measures

#### Anxiety symptoms

The 7 items General Anxiety Disorder (GAD-7) was used, and higher score indicating higher level of anxiety (Wang et al., 2014). An example items was, “feeling nervous, anxious or on edge”. In this study, the Cronbach’s alpha was 0.92, showing good internal reliability.

#### Interoceptive awareness

Three subscales (i.e., noticing, emotional awareness, and body listening) of Multidimensional Assessment of Interceptive Awareness (MAIA) were used to access participants’ awareness of body sensations (e.g., I notice when I am uncomfortable in my body),ability to attribute specific physical sensations to their psychological manifestations of emotions (e.g., I notice how my body changes when I am angry), tendency to actively listen to the body (e.g., I listen to my body to inform me about what to do; Lin et al., 2017 & Mehling et al., 2012). In the current study, these subscales demonstrated acceptable internal reliability, with a range of Cronbach’s alpha from 0.73 to 0.91.

#### NSSI behavior

The single item was used to measure individual engaged in NSSI behavior in their lifetime, for example, “how many times have you deliberately hurt yourself but without an intention to die” (Gandhi et al., 2018).

### Statistical analysis

In the first analysis, we first recorded the NSSI lifetime history into 0 = No experience and 1 = Had experience (*N* _Total number_ = 5281) following Gandhi’s previous work (Gandhi et al., 2018). The multilevel logistic regression (level 1: demographic information, including gender, age and subjective family income, Lindberg et al., 2021; level 2: emotional level; level 3: noticing, emotional awareness and body listening of interoceptive awareness) was used to examine the effects of these variables on the individual’s experience of NSSI behaviors.

In the second analysis, only participants with NSSI history were included (N=279). We then recorded the frequency of NSSI behavior into 0 = less or equal to five times and 1 = higher than five times. The Multiple mediator model and Bootstrap with 1000 times (Tofighi, 2020), was used to examine the indirect effect of Noticing, Emotional awareness and Body listening on the association between anxiety symptoms and NSSI behaviors. Jamovi 1.2.25 was used for data analysis. Statistical significance was set at *p*<.05.

## Results

The prevalence of lifetime NSSI behaviors among Chinese university student was 5.35% (N = 279/5002). As shown in Table 1, female (OR = 1.40, *p* = .02), younger (OR = -0.14, *p* <.001), a higher level of anxiety symptoms (OR = 1.11, *p* <.001), a higher level of Noticing (OR = 1.14, *p* <.001), a lower level of Emotion Awareness (OR = 0.95, *p* =.01), and a higher level of Body listening (OR = 1.27, *p* =.01) significantly predicted higher probability of individual’s NSSI behaviors.

**Table 1.** Multilevel Logistic Regression Modelling Results for the NSSI Behaviors (*N =* 5281).

|  | Model 1 |  |  |  | Model 2 |  |  |  | Model 3 |  |  |  |
| --- | --- | --- | --- | --- | --- | --- | --- | --- | --- | --- | --- | --- |
|  | *B* | *p* | OR | 95% C.I | *B* | *p* | OR | 95% C.I | *B* | *p* | OR | 95% C.I |
| **Intercept** | 0.80 | .35 | 2.23 | [0.41, 12.05] | -1.88 | .03 | 0.15 | [0.03, 0.87] | -4.50 | <.001 | 0.01 | [0.002,0.07] |
| Age | -0.19 | <.001 | 0.83 | [0.77, 0.89] | -0.17 | <.001 | 0.84 | [0.78, 0.91] | -0.14 | <.001 | 0.85 | [0.81, 0.94] |
| Gender | 0.36 | .01 | 0.83 | [1.09, 1.90] | 0.35 | .014 | 1.43 | [1.07, 1.89] | 0.33 | .023 | 1.40 | [1.05, 1.86] |
| Income | -0.07 | .04 | 0.93 | [0.87, 0.996] | -0.01 | .82 | 0.99 | [0.92, 1.07] | 0.003 | .94 | 1.00 | [0.93, 1.08] |
| Anxiety |  |  |  |  | 0.15 | <.001 | 1.16 | [1.13, 1.19] | 0.10 | <.001 | 1.11 | [1.08, 1.14] |
| Noticing |  |  |  |  |  |  |  |  | 0.13 | <.001 | 1.14 | [1.08, 1.20] |
| Awareness |  |  |  |  |  |  |  |  | -0.05 | 0.01 | 0.95 | [0.92, 0.99] |
| Listening |  |  |  |  |  |  |  |  | 0.24 | <.001 | 1.27 | [1.16, 1.39] |
| $\chi^2$ | | | | 42.7 | | | | 178.4 | | | | 249.4 |
| *p* |  |  |  | <.001 |  |  |  | <.001 |  |  |  | <.001 |

**Note.** Income = subjective family income, Anxiety = Anxiety symptoms, Noticing = The body noticing in interoceptive awareness, Listening = The body listening in interoceptive awareness, Awareness = The emotion awareness in interoceptive awareness. Female was coded as 2, and male was coded as 1.

The result of multiple mediator model is shown in Figure 1. The results showed that the direct effect of anxiety symptoms on NSSI behavior was not significant (β = 0.13, 95% CI [<-0.001, 0.02]), and the total effect was significant, with β =0.14, 95% CI [0.002, 0.02].

**Figure 1.**
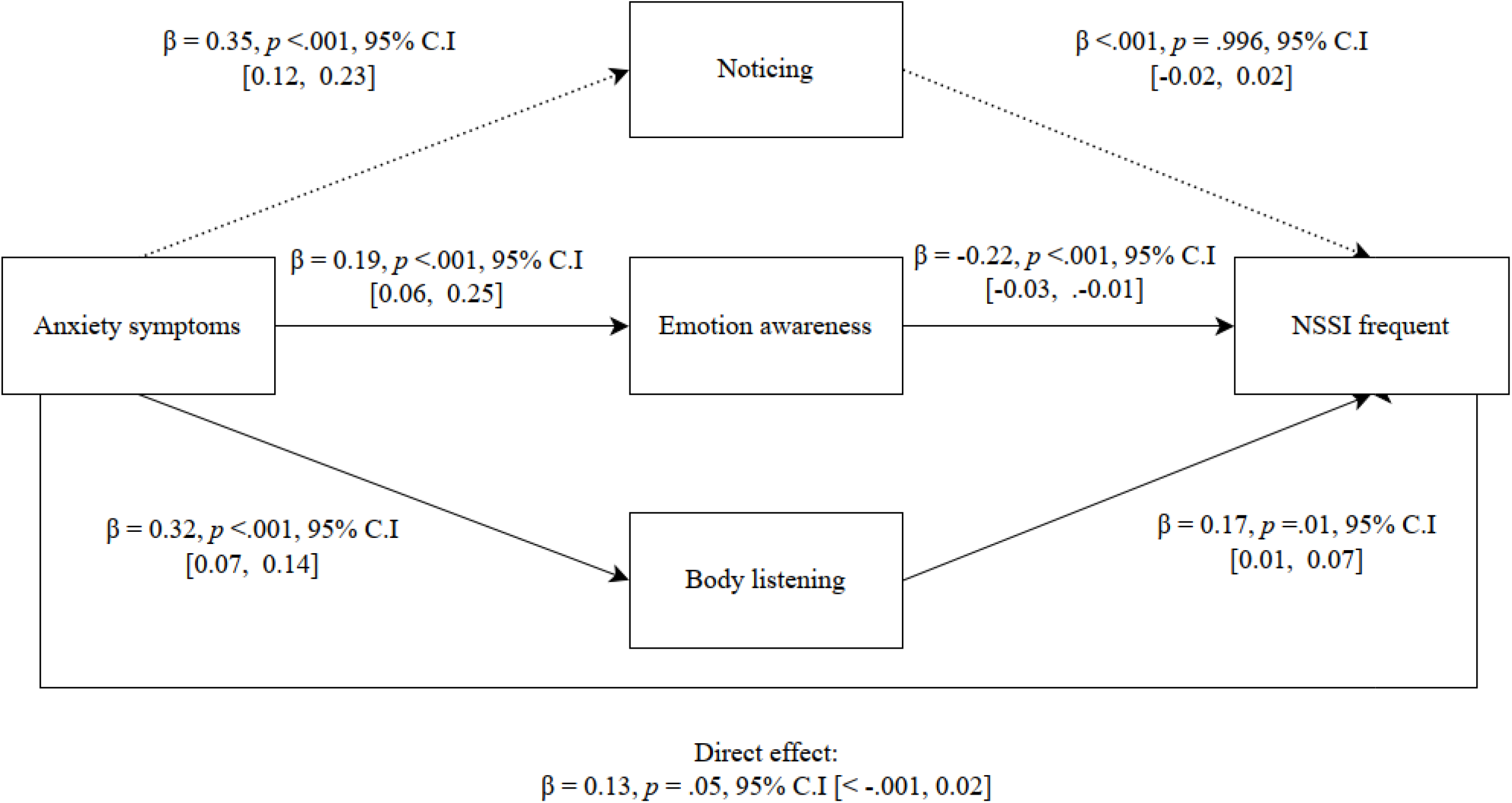
The multiple mediator model of the association between anxiety symptoms and NSSI frequent via, Noticing, Emotion awareness, and Body listening in interoceptive awareness. The outcome was standardized, and beta are completely standardized effect sizes.

Whereas, the effect of anxiety symptoms on Noticing (β = 0.35, 95% CI [0.12, 0.23]), Emotional awareness (β = 0.19, 95% CI [0.06, 0.25]) and Body listening (β = 0.32, 95% CI [0.07, 0.14]) was significant, as well as that of Emotional awareness (β = -0.22, 95% CI [-0.03, -0.01]) and Body listening (β = 0.17, 95% CI [0.01, 0.07]) on NSSI frequent. However, the effect of Noticing on NSSI frequent was not significant, β <.001, 95% CI [-0.02, 0.02].

In this model, the indirect effect of anxiety on NSSI behavior via Body listening and Emotional awareness were significant, β = 0.06, 95% CI [<0.001, 0.01], which accounted 43% for total variance; and β = -0.04, 95% CI [-0.01, <-0.001], respectively; whereas, the indirect pathway throughout the Noticing was not significant, β < -0.001, 95% CI [-0.003, 0.003].

## Discussion

In the present study, the prevalence of lifetime NSSI behavior among youth was reached 5.35%, which is lower than the prevalence found among 11 to 29 years youth in Scotland (i.e., 7.1%), and a meta-analysis focused on middle-school student in China (Lang & Yao, 2018).

We further found the younger and female showed higher probability of lifetime NSSI behaviors. These findings further warrant us the existing of heterogeneity (i.e., cultural diversity and age difference) among group’s NSSI behaviors, which may also be the reason for the difficulty to fully understand the underlying mechanisms with respect to NSSI. In this case, to the best of our knowledge, this is a very first study to clarify the potential pathways which contributes to NSSI behaviors by focusing on a special and importance stage of Chinese population.

Following the first analysis among general students, which found a higher level of anxiety symptoms, a higher level of Noticing, a lower level of Emotion Awareness, and a higher level of Body listening significantly predicted higher probability of individual’s NSSI behaviors; we then conducted a path analysis to further clarify these relationships among individual had NSSI behaviors. Interesting, Emotion awareness and Body listening plays an indirect effect between anxiety symptoms and lifetime NSSI behaviors, but in a opposite way. Specifically, in line with the integrate affective and cognitive processing model of anxiety disorders (Paulus & Stein, 2006) and several previous studies (Domschke et al., 2010; Dunn et al., 2010; Young et al., 2021), we found individual with higher level of anxiety symptoms tends to show higher or might be overly sensitive body listening abilities, this may further increase their possibility in taking higher frequent NSSI behaviors, as a subsequent dysfunctional emotional regulation strategies.

However, we amazingly found that the higher abilities of Emotion awareness which triggers by higher level of anxiety symptoms would decrease the possibilities of NSSI. This discrepancy may be due to the fact that the independence of interoceptive dimensions has been neglected in previous studies. It showing the fact that, Emotional awareness as a precondition of one’s abilities to regulate their emotion in adaptative way (Füstös et al., 2013); a strength of this abilities may further reduced one’s maladaptive emotion regulation strategies, that is, NSSI behaviors.

This novelty double-edged sword finding largely extend the previous theories on dimensionally view of interoceptive awareness (Khalsa et al., 2018; Mehling et al., 2018; Mehling et al., 2012), and how its transit between emotion distress to their maladaptive behaviors. For the clinical practice, these findings shed a light on the future preventive intervention program for the urgent in helping individual with anxiety symptoms to restoring their abilities of awareness for “what my body represent my feelings”, rather than simple be the “slaves of body”, which is consistent with the calling of body-mind approach (Burnett-Zeigler et al., 2016).

Though this study shows promise strategies for a more cost-effective interception-based intervention, it still had several limitations, including the cross-sectional and self-report design which may leading to bias (Adams et al., 1999; Rosenman et al., 2011); future studies are required to have a more objective and time-series design. In addition, the difference between trait anxiety and state anxiety also needs more clarifies (Leal et al., 2017).

## Data Availability

All data produced in the present study are available upon reasonable request to the authors

## References

Adams, A. S., Soumerai, S. B., Lomas, J., & Ross-Degnan, D. (1999). Evidence of self-report bias in assessing adherence to guidelines. International Journal for Quality in Health Care, 11(3), 187–192.

Bentley, K. H., Cassiello-Robbins, C. F., Vittorio, L., Sauer-Zavala, S., & Barlow, D. H. (2015). The association between nonsuicidal self-injury and the emotional disorders: A meta-analytic review. Clinical Psychology Review, 37, 72–88. https://doi.org/https://doi.org/10.1016/j.cpr.2015.02.006

Bock, R. C., Berghoff, C. R., Baker, L. D., Tull, M. T., & Gratz, K. L. (2021). The Relation of Anxiety to Nonsuicidal Self Injury Is Indirect Through Mindfulness. Mindfulness, 12(8), 2022–2033. https://doi.org/10.1007/s12671-021-01660-2

Burnett-Zeigler, I., Schuette, S., Victorson, D., & Wisner, K. L. (2016). Mind–body approaches to treating mental health symptoms among disadvantaged populations: A comprehensive review. The Journal of Alternative and Complementary Medicine, 22(2), 115–124.

Daukantaite, D., Lundh, L.-G., Wångby-Lundh, M., Claréus, B., Bjärehed, J., Zhou, Y., & Liljedahl, S. I. (2021). What happens to young adults who have engaged in self-injurious behavior as adolescents? A 10-year follow-up. European Child & Adolescent Psychiatry, 30(3), 475–492. https://doi.org/10.1007/s00787-020-01533-4

Domschke, K., Stevens, S., Pfleiderer, B., & Gerlach, A. L. (2010). Interoceptive sensitivity in anxiety and anxiety disorders: an overview and integration of neurobiological findings. Clinical Psychology Review, 30(1), 1–11.

Dunn, B. D., Stefanovitch, I., Evans, D., Oliver, C., Hawkins, A., & Dalgleish, T. (2010). Can you feel the beat? Interoceptive awareness is an interactive function of anxiety- and depression-specific symptom dimensions. Behaviour Research and Therapy, 48(11), 1133–1138. https://doi.org/https://doi.org/10.1016/j.brat.2010.07.006

Füstös, J., Gramann, K., Herbert, B. M., & Pollatos, O. (2013). On the embodiment of emotion regulation: interoceptive awareness facilitates reappraisal. Social Cognitive and Affective Neuroscience, 8(8), 911–917. https://doi.org/10.1093/scan/nss089

Gandhi, A., Luyckx, K., Goossens, L., Maitra, S., & Claes, L. (2018). Association between Non-Suicidal Self-Injury, Parents and Peers Related Loneliness, and Attitude Towards Aloneness in Flemish Adolescents: An Empirical Note. Psychologica Belgica, 58, 3–12. https://doi.org/10.5334/pb.385

Garfinkel, S., Critchley, H., & Pollatos, O. (2015). The interoceptive system: implications for cognition, emotion, and health. In Handbook of psychophysiology (pp. 427-443). Cambridge University Press.

Garisch, J. A., & Wilson, M. S. (2015). Prevalence, correlates, and prospective predictors of non-suicidal self-injury among New Zealand adolescents: cross-sectional and longitudinal survey data. Child and Adolescent Psychiatry and Mental Health, 9(1), 28. https://doi.org/10.1186/s13034-015-0055-6

Hasking, P., Whitlock, J., Voon, D., & Rose, A. (2017). A cognitive-emotional model of NSSI: using emotion regulation and cognitive processes to explain why people self-injure. Cognition and Emotion, 31(8), 1543–1556. https://doi.org/10.1080/02699931.2016.1241219

Khalsa, S. S., Adolphs, R., Cameron, O. G., Critchley, H. D., Davenport, P. W., Feinstein, J. S., Feusner, J. D., Garfinkel, S. N., Lane, R. D., & Mehling, W. E. (2018). Interoception and mental health: a roadmap. Biological psychiatry: cognitive neuroscience and neuroimaging, 3(6), 501–513.

Lang, J., & Yao, Y. (2018). Prevalence of nonsuicidal self-injury in chinese middle school and high school students: A meta-analysis. Medicine, 97(42), e12916–e12916. https://doi.org/10.1097/MD.0000000000012916

Leal, P. C., Goes, T. C., da Silva, L. C. F., & Teixeira-Silva, F. (2017). Trait vs. state anxiety in different threatening situations. Trends in psychiatry and psychotherapy, 39, 147–157.

Lindberg, M. H., Chen, G., Olsen, J. A., & Abelsen, B. (2021). Explaining subjective social status in two countries: The relative importance of education, occupation, income and childhood circumstances. SSM - Population Health, 15, 100864. https://doi.org/https://doi.org/10.1016/j.ssmph.2021.100864

Lundh, L.-G., Wångby-Lundh, M., Paaske, M., Ingesson, S., & Bjärehed, J. (2011). Depressive symptoms and deliberate self-harm in a community sample of adolescents: a prospective study. Depression research and treatment, 2011.

Mehling, W. E., Acree, M., Stewart, A., Silas, J., & Jones, A. (2018). The Multidimensional Assessment of Interoceptive Awareness, Version 2 (MAIA-2). PLoS One, 13(12), e0208034–e0208034. https://doi.org/10.1371/journal.pone.0208034

Mehling, W. E., Price, C., Daubenmier, J. J., Acree, M., Bartmess, E., & Stewart, A. (2012). The Multidimensional Assessment of Interoceptive Awareness (MAIA). PLoS One, 7(11), e48230. https://doi.org/10.1371/journal.pone.0048230

Paulus, M. P., & Stein, M. B. (2006). An Insular View of Anxiety. Biological Psychiatry, 60(4), 383–387. https://doi.org/10.1016/j.biopsych.2006.03.042

Rosenman, R., Tennekoon, V., & Hill, L. G. (2011). Measuring bias in self-reported data. International Journal of Behavioural and Healthcare Research, 2(4), 320–332.

Taylor, P. J., Jomar, K., Dhingra, K., Forrester, R., Shahmalak, U., & Dickson, J. M. (2018). A meta-analysis of the prevalence of different functions of non-suicidal self-injury. Journal of Affective Disorders, 227, 759–769. https://doi.org/https://doi.org/10.1016/j.jad.2017.11.073

Tofighi, D. (2020). Bootstrap Model-Based Constrained Optimization Tests of Indirect Effects [Methods]. Frontiers in Psychology, 10. https://doi.org/10.3389/fpsyg.2019.02989

Wang, W., Bian, Q., Zhao, Y., Li, X., Wang, W., Du, J., Zhang, G., Zhou, Q., & Zhao, M. (2014). Reliability and validity of the Chinese version of the Patient Health Questionnaire (PHQ-9) in the general population. General Hospital Psychiatry, 36. https://doi.org/10.1016/j.genhosppsych.2014.05.021

Wolff, J. C., Thompson, E., Thomas, S. A., Nesi, J., Bettis, A. H., Ransford, B., Scopelliti, K., Frazier, E. A., & Liu, R. T. (2019). Emotion dysregulation and non-suicidal self-injury: A systematic review and meta-analysis. Eur Psychiatry, 59, 25–36. https://doi.org/10.1016/j.eurpsy.2019.03.004

Young, H. A., Davies, J., Freegard, G., & Benton, D. (2021). Nonsuicidal self-injury is associated with attenuated interoceptive responses to self-critical rumination. Behavior Therapy, 52(5), 1123–1136.

